# Metabolite Signature of Life’s Essential 8 and Risk of Coronary Heart Disease among Low-Income Black and White Americans

**DOI:** 10.1101/2023.04.24.23289055

**Authors:** Kui Deng, Deepak K. Gupta, Xiao-Ou Shu, Loren Lipworth, Wei Zheng, Victoria E. Thomas, Hui Cai, Qiuyin Cai, Thomas J. Wang, Danxia Yu

**Author notes:** Correspondence: Danxia Yu, Division of Epidemiology, Vanderbilt University Medical Center, 2525 West End Avenue, Nashville, TN, USA 37203.

## Abstract

**Background and Aims:** Life’s Essential 8 (LE8) is a comprehensive construct of cardiovascular health. Yet, little is known about LE8 score, its metabolic correlates, and their predictive implications among Black Americans and low-income individuals.

**Methods:** In a nested case-control study of coronary heart disease (CHD) among 598 Black and 596 White low-income Americans, we estimated LE8 score, conducted untargeted plasma metabolites profiling, and used elastic net with leave-one-out cross-validation to identify metabolite signature (MetaSig) of LE8. Associations of LE8 score and MetaSig with incident CHD were examined using conditional logistic regression. Mediation effect of MetaSig on the LE8-CHD association was also examined. The external validity of MetaSig was evaluated in another nested CHD case-control study among 598 Chinese adults.

**Results:** Higher LE8 score was associated with lower CHD risk [standardized OR (95% CI)=0.61 (0.53-0.69)]. The identified MetaSig, consisting of 133 metabolites, showed strong correlation with LE8 score (*r*=0.61) and inverse association with CHD risk [OR (95% CI)=0.57 (0.49-0.65)], robust to adjustment for LE8 score and across participants with different sociodemographic and health status (ORs: 0.42-0.69; all *P*<0.05). MetaSig mediated a large portion of the LE8-CHD association: 53% (32%-80%) (*P*<0.001). Significant associations of MetaSig with LE8 score and CHD risk were found in validation cohort [*r*=0.49; OR (95% CI)=0.57 (0.46-0.69)].

**Conclusions:** Higher LE8 score and its MetaSig were associated with lower CHD risk among low-income Black and White Americans. Metabolomics may offer an objective and comprehensive measure of LE8 score and its metabolic phenotype relevant to CHD prevention among diverse populations.

## Introduction

Coronary heart disease (CHD) is a leading cause of morbidity and mortality in the United States (US) and worldwide, with significant and persistent sociodemographic disparities^1, 2^. To reduce the burden and life lost due to CHD and other cardiovascular disease (CVD), the American Heart Association (AHA) has recently proposed Life’s Essential 8 (LE8) to assess and promote cardiovascular health (CVH) in individuals and populations^3^. LE8 includes 4 health behaviors (healthy diet, participation in physical activity, avoidance of nicotine, and healthy sleep [a new component of LE8]) and 4 health factors (weight, blood lipids, glucose, and blood pressure), and has a new scoring system with continuous scale to better reflect inter-individual differences. While a higher LE8 score has been recently associated with lower CVD incidence and mortality^4–7^, few studies have evaluated LE8 score and its association with incident CVD among Black and White Americans who have low socioeconomic status (SES) and face disproportionate CVD burdens. In addition, although some potential mechanisms have been identified (e.g., reduced inflammation and atherosclerosis)^8, 9^, beyond those known CVD risk pathways, mechanisms and inter-individual differences underlying the cardioprotective effects of LE8 and its included health behaviors and health factors are not fully understood.

Metabolite profiling (“metabolomics”) comprehensively measures small-molecule metabolites (including substrates, intermediates, and end products) in biological samples and represents a powerful tool for mechanistic investigation, novel biomarker discovery, and precision medicine^10, 11^. Metabolite profiling of blood samples may improve assessments of individuals’ alignment with LE8, particularly for behavioral factors that are prone to survey and recall biases. In addition, circulating metabolites related to LE8 may capture varied individual metabolic responses to LE8, providing novel mechanistic insights into its cardioprotective effects and informing precision medicine. While previous studies have identified metabolites related to the components of LE8, including diet^12–16^, physical activity^17–19^, tobacco exposure^20–22^, sleep^23–26^, and body mass index (BMI)^27, 28^, to our knowledge, no study has applied untargeted plasma metabolomics to identify a comprehensive metabolite signature (MetaSig) for LE8 to enable studies with incident CHD. Given that those health behaviors and factors often correlate and interact with each other, investigating whether plasma metabolomics could provide a good objective assessment of individuals’ alignment with and metabolic responses to overall LE8 and uncovering potential pathways linking LE8 to incident CHD is highly warranted.

Here, leveraging a case-control study of CHD nested within the Southern Community Cohort Study (SCCS) involving 598 Black Americans and 596 White Americans, we assessed the LE8 score and its MetaSig and examined their relations to incident CHD. The results were further replicated in another nested CHD case-control study of racially and geographically different population: 598 Chinese adults from the Shanghai Women’s Health Study and Shanghai Men’s Health Studies (SWMHS). In addition, we identified MetaSigs for the health behaviors and health factors recommended in the LE8 and evaluated their associations with incident CHD.

## Methods

### Study participants

This study was based on a nested case-control study of CHD within the SCCS^29^. Briefly, the SCCS enrolled 84,735 primarily low-income, uninsured/underinsured Black and White Americans aged 40-79 years from 12 southeastern US states between 2002-2009, with >50% having household income less than $15,000/y and ∼86% were uninsured or underinsured^29^. Participants were surveyed for a wide range of information at baseline and followed up regularly for morbidity and mortality outcomes. Venous blood samples were collected at baseline, and plasma samples were aliquoted and stored at -80°C for long-term use. A nested case-control study of CHD within SWMHS was used as the validation cohort, which enrolled 74,940 women, aged 40-70 years, and 61,480 men, aged 40-74 years, from Shanghai, China between 1996–2000 and 2002–2006, respectively^30, 31^. These cohort studies were approved by the Institutional Review Boards of the Vanderbilt University Medical Center, Meharry Medical College, and/or Shanghai Cancer Institute. Informed consent was obtained from all enrolled participants.

For the nested case-control studies, participant inclusion criteria were 1) no history of CHD, stroke, heart failure, cancer, or end-stage renal disease at baseline; 2) available baseline plasma samples and data on fasting time and in SCCS, time between sample collection and lab processing; 3) no use of antibiotics nor cold/flu in last 7 days before blood collection. In addition, to facilitate CHD case identification and adjudication, in SCCS, participants were eligible for Centers for Medicare & Medicaid Services (CMS) and had ≥2 claims after cohort enrollment; in SWMHS, participants’ medical records were accessible for our study. In SCCS, nonfatal CHD cases were identified through CMS, including acute myocardial infarction, coronary revascularization, and other acute CHD, and CHD deaths were identified through the National Death Index. In SWMHS, CHD cases were first identified by self-reported diagnoses during follow-up visits and then confirmed by medical records. In each race (Black, White, or Chinese) and gender (male or female), 150 incident CHD cases were randomly selected and 1:1 matched with controls who had no CHD, heart failure, stroke, nor cancer at the time of case diagnosis, by enrollment age (±2 years), fasting time (±2 hours), and time between sample collection and lab processing (±4 hours, for SCCS samples; all SWMHS samples were processed within 6 hours after collection). After excluding eight plasma samples that did not pass the metabolite profiling quality control, a total of 1792 participants, including 597 pairs of CHD cases and controls in SCCS (299 pairs of Black Americans and 298 pairs of White Americans), and 299 pairs of Chinese adults in SWMHS were included in the present study (**Fig. 1**).

**Fig. 1.**
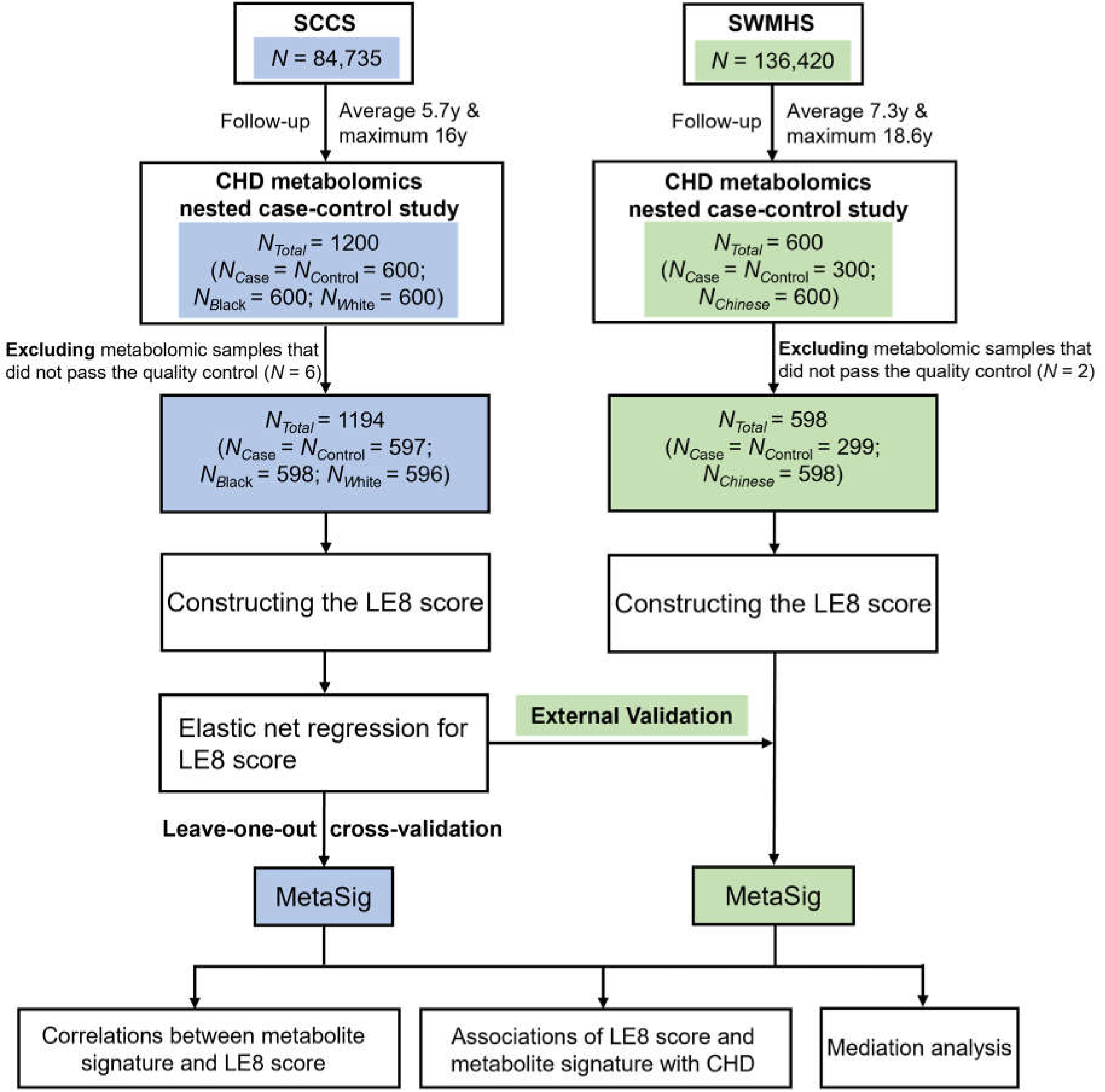
Overview of the current study. This study involved two nested case-control metabolomics studies within Southern Community Cohort Study (SCCS, primary cohort) and Shanghai Women’s and Men’s Health Studies (SWMHS, validation cohort). In the follow-up visits, participants with incident coronary heart disease (CHD) were identified and matched with controls by enrollment age, sex, race, fasting time, and time between sample collection and lab processing. After excluding 8 samples that did not pass metabolomics quality control, 1194 SCCS participants and 598 SWMHS participants were included. Metabolite signature of Life’s Essential 8 (LE8) were identified using elastic net regression with leave-one-out cross-validation in SCCS. The associations of LE8 score and its metabolite signature with risk of CHD were evaluated by conditional logistic regression adjusted for confounders. Mediation analysis was performed to assess the potential mediating role of metabolite signature on the LE8-CHD association. External validity of the identified metabolite signature related to LE8 score and CHD risk was investigated in SWMHS. **Abbreviations:** SCCS, Southern Community Cohort Study; SWMHS, Shanghai Women’s and Men’s Health Studies; CHD, coronary heart disease; LE8, Life’s Essential 8; MetaSig, metabolite signature.

### Life’s Essential 8

We constructed each LE8 component metric and total LE8 score (ranging from 0 to 100) according to AHA guidelines^3, 32^, with some modifications based on the characteristics of our data, as shown in **Table S1**. Briefly, dietary quality was assessed by the Dietary Approaches to Stop Hypertension (DASH) score^33–35^. Physical activity was measured by total minutes of leisure-time moderate and vigorous physical activity per week, with each minute of vigorous physical activity counted as 2 minutes toward the total minutes. Nicotine exposure was assessed by active tobacco smoking and secondhand smoking exposure. Sleep health was measured by average self-reported sleep hours per day. BMI was calculated as weight (kg)/height (m)^2^. Blood lipids component was assessed by non-HDL cholesterol (mg/dL) (plasma total cholesterol minus HDL cholesterol). Because concentrations of fasting glucose and HbA1c were not measured in our study, blood glucose component was scored based on diabetes diagnosis and medications and relative abundance of plasma glucose measured simultaneously with other metabolites (see following metabolite profiling methods). Blood pressure was unavailable in SCCS, thus blood pressure component was scored based on hypertension status and medications. Total LE8 score was obtained by calculating the arithmetic mean of individual component scores. We also calculated scores reflecting alignment with health behaviors and health factors.

### Metabolite profiling

Baseline plasma samples of selected CHD case-control pairs were retrieved and placed adjacently in the same assay batch. Laboratory persons were blinded to the case-control status of samples. Untargeted metabolite profiling was performed using ultra-high-performance liquid chromatography (UHPLC) coupled with tandem mass spectrometry (MS) by Metabolon Inc. (Morrisville, NC, USA) following a standard assay protocol^36^. Briefly, plasma samples were extracted with methanol and split into four aliquots for analysis by UHPLC-MS/MS in both positive and negative ion modes using a combination reverse phase and HILIC chromatography methods. Metabolites were identified by automated comparison of mass spectra features to a reference library of >4,000 authenticated standard compounds followed by visual inspection for quality control. Peaks were quantified using area-under-the-curve. A total of 1502 metabolites were detected in our samples. The majority of metabolites (>80%) were annotated based on internal standards. Metabolites that were annotated only by a match to a known MS spectrum or chemical formula were marked by ‘*’ and ‘**’, respectively. We excluded metabolites detected in <10% of participants, resulting in 1322 metabolites. Metabolites with missing values were imputed by half of the minimal value in the non-missing samples. The values of all metabolites were log-transformed and standardized to mean 0 and unit variance.

### Statistical analysis

The characteristics of the study participants were presented as mean (standard deviation [SD]) for continuous variables and frequency (percentage) for categorical variables. *Spearman* correlations between LE8 score and individual component scores were assessed. Elastic net regression was used to identify metabolites associated with LE8^12, 14, 37^, with hyperparameters determined via a ten-repeated tenfold cross-validation framework, using R package caret (version: 6.0-88)^38^ and glmnet (version: 4.1-3)^37, 39^. The MetaSig was calculated through a leave-one-out cross-validation approach. In addition, we externally validated the identified signature in SWMHS using weights of the selected metabolites from SCCS. We calculated *Spearman* correlations between LE8 score and MetaSig among all participants and by race, sex, age (≥60y/<60y), incident CHD status, diabetes status, hypertension status, dyslipidemia status, and fasting status.

We then examined the associations of LE8 and its MetaSig with risk of CHD using conditional logistic regression, adjusting for age, education, income, alcohol intake, and family history of CHD. We also included LE8 score and MetaSig in the same model to assess their independent associations with CHD and potential mediating effect of MetaSig on LE8-CHD association. The potential multicollinearity was assessed by variance inflation factor (VIF) using R package car (version: 3.1-0) with VIF >10 indicating multicollinearity among variables. The ranges of VIF for LE8 score and its MetaSig were 1.08-1.37, confirming no multicollinearity. The mediation analysis was performed using R package mediation (version: 4.5.0)^40^. Subgroup analyses to evaluate the associations of LE8 and MetaSig with incident CHD were performed by race, age group, sex, education, household income, diabetes status, hypertension status, and dyslipidemia status, with *P* value for interaction obtained from the corresponding interaction term in the model.

The same methods were used to identify MetaSigs for health behaviors and health factors included in LE8 and evaluate their associations with incident CHD, potential mediation effects, and their external validity. Mutual adjustments of health behaviors and health factors score were performed in corresponding statistical models. All analyses were performed using R (version 4.1.1). Two-sided *P* < 0.05 was considered statistically significant. An overview of our study design is presented in **Fig. 1**.

## Results

### Baseline characteristics of study participants

The mean age at baseline (blood collection) was 55 years in our study participants (**Table 1**). The mean (SD) of LE8 score was 48.1 (12.3) in Black women, 50.0 (13.6) in Black men, 48.1 (13.4) in White women, and 48.5 (14.4) in White men. The median follow-up time for incident CHD cases was 5 (interquartile range: 3-8) years in SCCS. Incident CHD cases had significantly lower total LE8 score, health behaviors score, and health factors score than controls (all *P* <0.05). The characteristics of participants in SWMHS (mean age: 61 years; mean LE8 score: 57.2 in women and 50.7 in men) are shown in **Table S2**. There were moderate correlations between total LE8 score and individual component scores (*r* ranged from 0.22 with smoking to 0.47 with BMI and blood pressure scores in SCCS; **Fig. S1**).

**Table 1.**
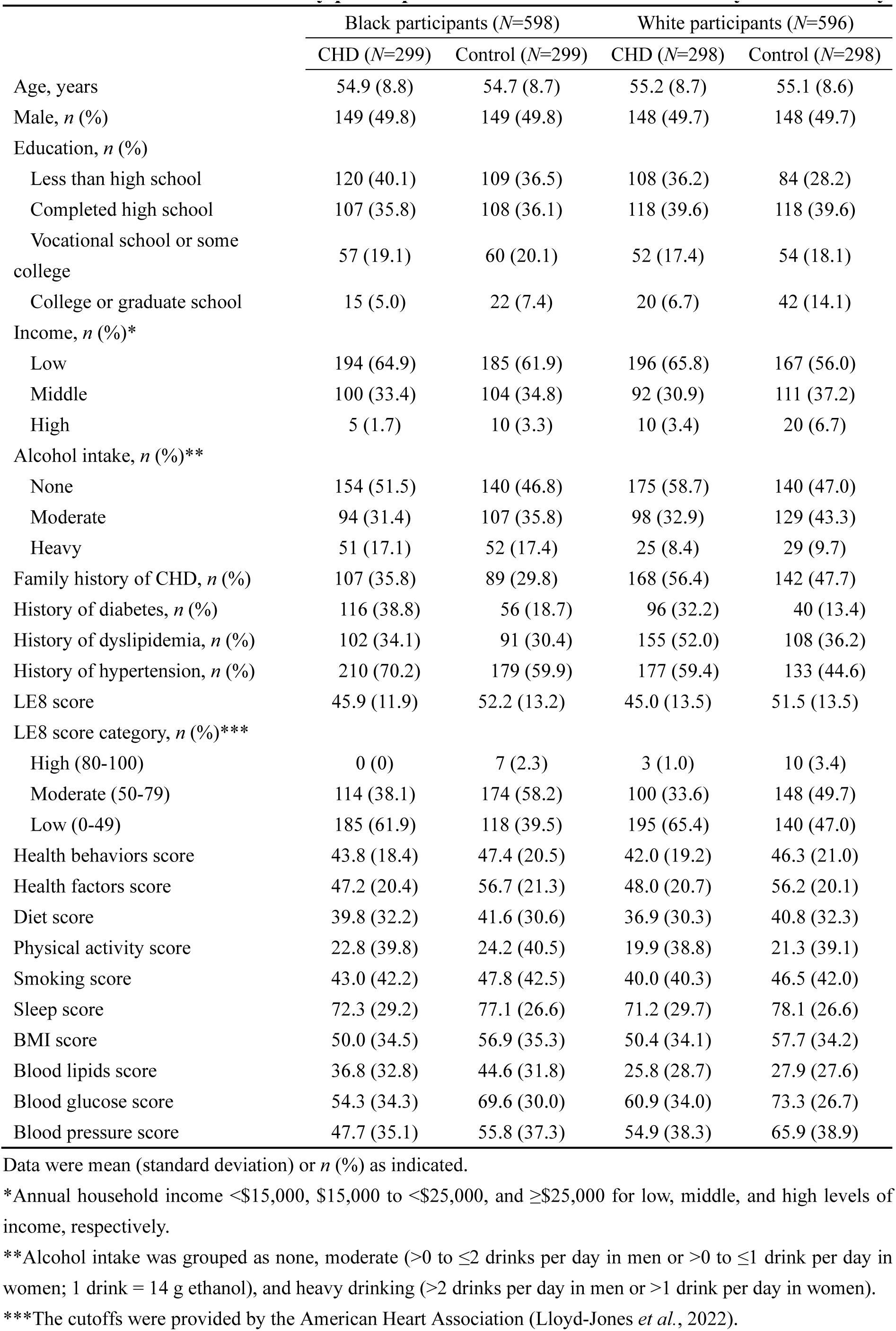
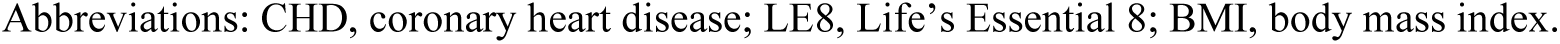
Characteristics of study participants in the Southern Community Cohort Study.

|  | Black participants (N=598) |  | White participants (N=596) |  |
| --- | --- | --- | --- | --- |
|  | CHD (N=299) | Control (N=299) | CHD (N=298) | Control (N=298) |
| Age, years | 54.9 (8.8) | 54.7 (8.7) | 55.2 (8.7) | 55.1 (8.6) |
| Male, *n* (%) | 149 (49.8) | 149 (49.8) | 148 (49.7) | 148 (49.7) |
| Education, *n* (%) |  |  |  |  |
| Less than high school | 120 (40.1) | 109 (36.5) | 108 (36.2) | 84 (28.2) |
| Completed high school | 107 (35.8) | 108 (36.1) | 118 (39.6) | 118 (39.6) |
| Vocational school or some college | 57 (19.1) | 60 (20.1) | 52 (17.4) | 54 (18.1) |
| College or graduate school | 15 (5.0) | 22 (7.4) | 20 (6.7) | 42 (14.1) |
| Income, *n* (%)* |  |  |  |  |
| Low | 194 (64.9) | 185 (61.9) | 196 (65.8) | 167 (56.0) |
| Middle | 100 (33.4) | 104 (34.8) | 92 (30.9) | 111 (37.2) |
| High | 5 (1.7) | 10 (3.3) | 10 (3.4) | 20 (6.7) |
| Alcohol intake, *n* (%)** |  |  |  |  |
| None | 154 (51.5) | 140 (46.8) | 175 (58.7) | 140 (47.0) |
| Moderate | 94 (31.4) | 107 (35.8) | 98 (32.9) | 129 (43.3) |
| Heavy | 51 (17.1) | 52 (17.4) | 25 (8.4) | 29 (9.7) |
| Family history of CHD, *n* (%) | 107 (35.8) | 89 (29.8) | 168 (56.4) | 142 (47.7) |
| History of diabetes, *n* (%) | 116 (38.8) | 56 (18.7) | 96 (32.2) | 40 (13.4) |
| History of dyslipidemia, *n* (%) | 102 (34.1) | 91 (30.4) | 155 (52.0) | 108 (36.2) |
| History of hypertension, *n* (%) | 210 (70.2) | 179 (59.9) | 177 (59.4) | 133 (44.6) |
| LE8 score | 45.9 (11.9) | 52.2 (13.2) | 45.0 (13.5) | 51.5 (13.5) |
| LE8 score category, *n* (%)*** |  |  |  |  |
| High (80-100) | 0 (0) | 7 (2.3) | 3 (1.0) | 10 (3.4) |
| Moderate (50-79) | 114 (38.1) | 174 (58.2) | 100 (33.6) | 148 (49.7) |
| Low (0-49) | 185 (61.9) | 118 (39.5) | 195 (65.4) | 140 (47.0) |
| Health behaviors score | 43.8 (18.4) | 47.4 (20.5) | 42.0 (19.2) | 46.3 (21.0) |
| Health factors score | 47.2 (20.4) | 56.7 (21.3) | 48.0 (20.7) | 56.2 (20.1) |
| Diet score | 39.8 (32.2) | 41.6 (30.6) | 36.9 (30.3) | 40.8 (32.3) |
| Physical activity score | 22.8 (39.8) | 24.2 (40.5) | 19.9 (38.8) | 21.3 (39.1) |
| Smoking score | 43.0 (42.2) | 47.8 (42.5) | 40.0 (40.3) | 46.5 (42.0) |
| Sleep score | 72.3 (29.2) | 77.1 (26.6) | 71.2 (29.7) | 78.1 (26.6) |
| BMI score | 50.0 (34.5) | 56.9 (35.3) | 50.4 (34.1) | 57.7 (34.2) |
| Blood lipids score | 36.8 (32.8) | 44.6 (31.8) | 25.8 (28.7) | 27.9 (27.6) |
| Blood glucose score | 54.3 (34.3) | 69.6 (30.0) | 60.9 (34.0) | 73.3 (26.7) |
| Blood pressure score | 47.7 (35.1) | 55.8 (37.3) | 54.9 (38.3) | 65.9 (38.9) |

Data were mean (standard deviation) or *n* (%) as indicated.
\*Annual household income <\$15,000, \$15,000 to <\$25,000, and ≥\$25,000 for low, middle, and high levels of income, respectively.
\*\*Alcohol intake was grouped as none, moderate (>0 to ≤2 drinks per day in men or >0 to ≤1 drink per day in women; 1 drink = 14 g ethanol), and heavy drinking (>2 drinks per day in men or >1 drink per day in women).
\*\*\*The cutoffs were provided by the American Heart Association (Lloyd-Jones *et al.*, 2022).
Abbreviations: CHD, coronary heart disease; LE8, Life's Essential 8; BMI, body mass index.

### Metabolite signature of LE8

We identified 133 metabolites related to LE8 (top 30 are shown in **Fig. 2A**; the full list can be found in **Table S3**). The MetaSig was strongly correlated with LE8 score (*r* = 0.61, *P*<0.001; **Fig. 2B**), meanwhile variations in MetaSig were observed among individuals with the same LE8 score, demonstrating interindividual difference in metabolic phenotype of LE8. MetaSig was also correlated with LE8 score in SWMHS (*r* = 0.49, *P*<0.001; **Fig. 2C**). Stratified analyses showed that correlations between LE8 score and MetaSig were consistent regardless of age, sex, race, fasting status, diabetes status, hypertension status, dyslipidemia status, and incident CHD status (*r* ranged from 0.54 to 0.64; **Table S4**), suggesting the robustness of our identified LE8 metabolite signature across participants with different sociodemographic backgrounds and metabolic disease status.

**Fig. 2.**
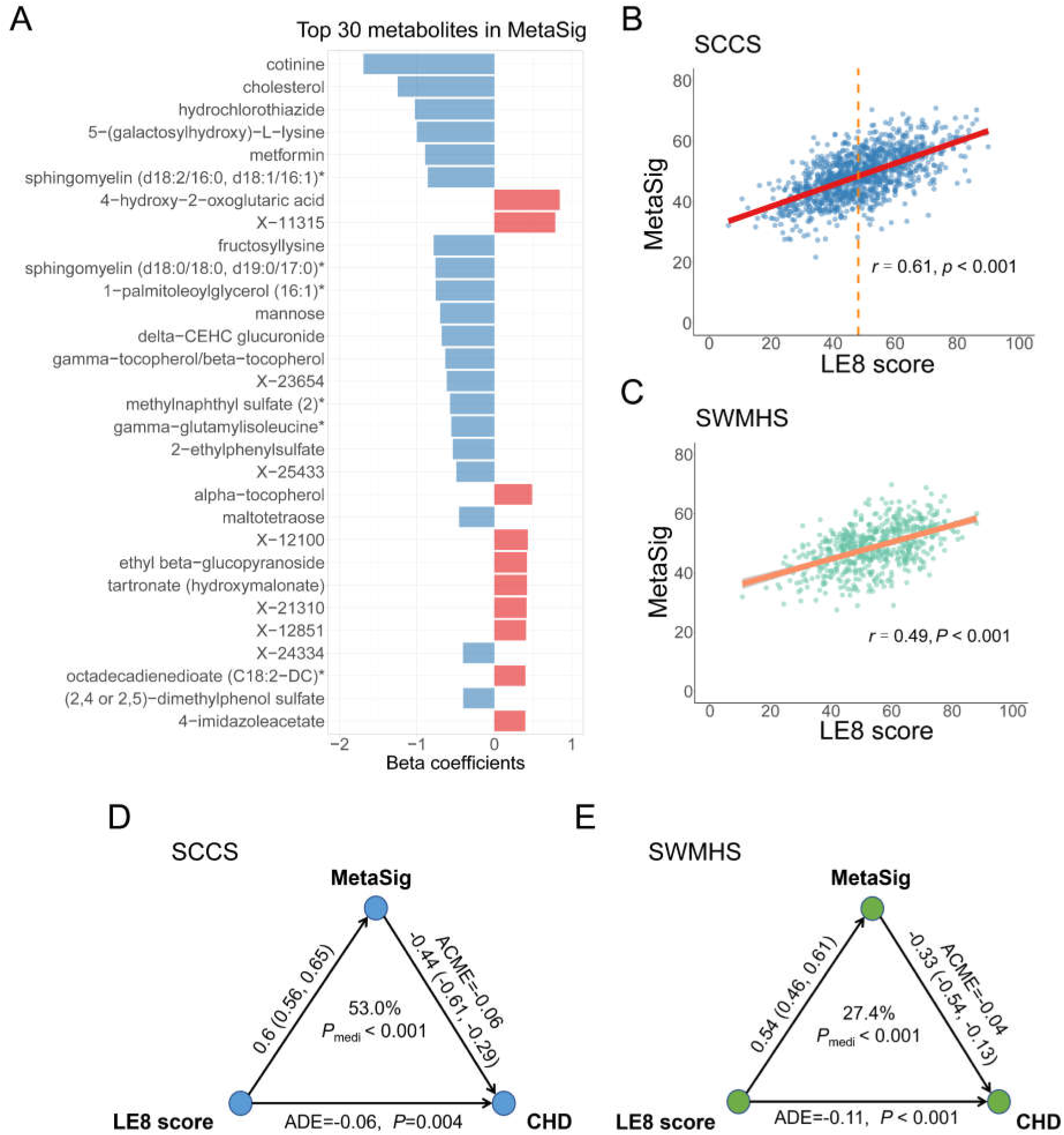
The metabolite signature of LE8 and its association with risk of CHD. **(A)** Top 30 metabolites selected by elastic net regression in SCCS. Metabolites were ranked by the absolute value of regression coefficients. **(B)** *Spearman* correlation between MetaSig and LE8 score in SCCS. The dashed line denotes median LE8 score. **(C)** *Spearman* correlation between MetaSig and LE8 score in SWMHS. **(D)** The mediation effect of MetaSig on the association between LE8 score and risk of CHD in SCCS. **(E)** The mediation effect of MetaSig on the association between LE8 score and risk of CHD in SWMHS. **Abbreviations:** SCCS, Southern Community Cohort Study; SWMHS, Shanghai Women’s and Men’s Health Studies; LE8, Life’s Essential 8; MetaSig, metabolite signature; ACME, average causal mediation effects; ADE, average direct effects; CHD, coronary heart disease.

### Associations with incident CHD

Higher LE8 score and its MetaSig were significantly associated with lower risk of CHD: standardized multivariable-adjusted odds ratio (OR) = 0.61 (95% CI: 0.53-0.69) for LE8 score and 0.57 (0.49-0.65) for MetaSig; both *P* < 0.001 (**Table 2**). Sensitivity analysis showed that the MetaSig-CHD associations did not change after excluding any individual metabolites from the signature (**Table S5**). After further adjusting for LE8 score, the MetaSig-CHD association was only slightly attenuated [OR (95% CI) = 0.66 (0.55-0.78); *P*<0.001; **Table 2**], suggesting circulating metabolites may complement LE8 assessment and contribute to CHD risk beyond LE8 score. On the other hand, the LE8-CHD association was moderately attenuated after adjusting for MetaSig [OR (95% CI) = 0.78 (0.66-0.92), *P*=0.003]. Mediation analysis showed that MetaSig mediated a large portion of the LE8-CHD association [53% (32%-80%); *P*_mediation_<0.001; **Fig. 2D**].

**Table 2.**
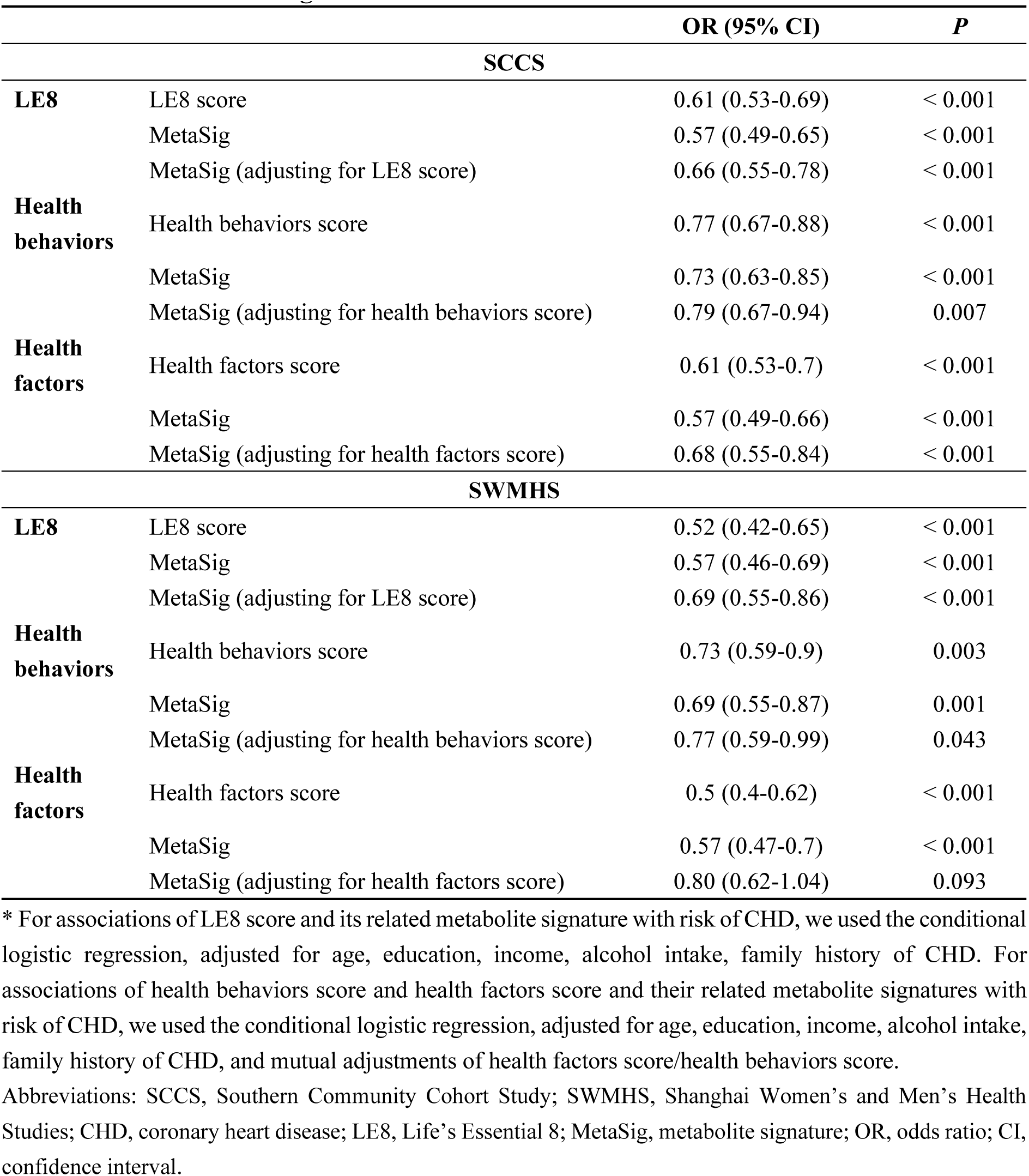
The associations of LE8 score, health behaviors score, health factors score and their related metabolite signatures with risk of CHD.

|  |  | OR (95% CI) | P |
| --- | --- | --- | --- |
| SCCS |  |  |  |
| **LE8** | LE8 score | 0.61 (0.53-0.69) | < 0.001 |
|  | MetaSig | 0.57 (0.49-0.65) | < 0.001 |
|  | MetaSig (adjusting for LE8 score) | 0.66 (0.55-0.78) | < 0.001 |
| **Health behaviors** | Health behaviors score | 0.77 (0.67-0.88) | < 0.001 |
|  | MetaSig | 0.73 (0.63-0.85) | < 0.001 |
|  | MetaSig (adjusting for health behaviors score) | 0.79 (0.67-0.94) | 0.007 |
| **Health factors** | Health factors score | 0.61 (0.53-0.7) | < 0.001 |
|  | MetaSig | 0.57 (0.49-0.66) | < 0.001 |
|  | MetaSig (adjusting for health factors score) | 0.68 (0.55-0.84) | < 0.001 |
| SWMHS |  |  |  |
| **LE8** | LE8 score | 0.52 (0.42-0.65) | < 0.001 |
|  | MetaSig | 0.57 (0.46-0.69) | < 0.001 |
|  | MetaSig (adjusting for LE8 score) | 0.69 (0.55-0.86) | < 0.001 |
| **Health behaviors** | Health behaviors score | 0.73 (0.59-0.9) | 0.003 |
|  | MetaSig | 0.69 (0.55-0.87) | 0.001 |
|  | MetaSig (adjusting for health behaviors score) | 0.77 (0.59-0.99) | 0.043 |
| **Health factors** | Health factors score | 0.5 (0.4-0.62) | < 0.001 |
|  | MetaSig | 0.57 (0.47-0.7) | < 0.001 |
|  | MetaSig (adjusting for health factors score) | 0.80 (0.62-1.04) | 0.093 |

\* For associations of LE8 score and its related metabolite signature with risk of CHD, we used the conditional logistic regression, adjusted for age, education, income, alcohol intake, family history of CHD. For associations of health behaviors score and health factors score and their related metabolite signatures with risk of CHD, we used the conditional logistic regression, adjusted for age, education, income, alcohol intake, family history of CHD, and mutual adjustments of health factors score/health behaviors score. Abbreviations: SCCS, Southern Community Cohort Study; SWMHS, Shanghai Women's and Men's Health Studies; CHD, coronary heart disease; LE8, Life's Essential 8; MetaSig, metabolite signature; OR, odds ratio; CI, confidence interval.

Both LE8 and its MetaSig were inversely associated with CHD risk in subpopulations by race, age group, education, income, diabetes status, hypertension status, and dyslipidemia status (*P*_interaction_>0.05; **Fig. 3**), with stronger associations observed in women than in men [for LE8, OR (95% CI) = 0.53 (0.42-0.66) in women and 0.66 (0.55-0.80) in men, *P*_interaction_ = 0.017; for MetaSig, 0.48 (0.38-0.6) in women and 0.65 (0.54-0.78) in men, *P*_interaction_ = 0.016].

**Fig. 3.**
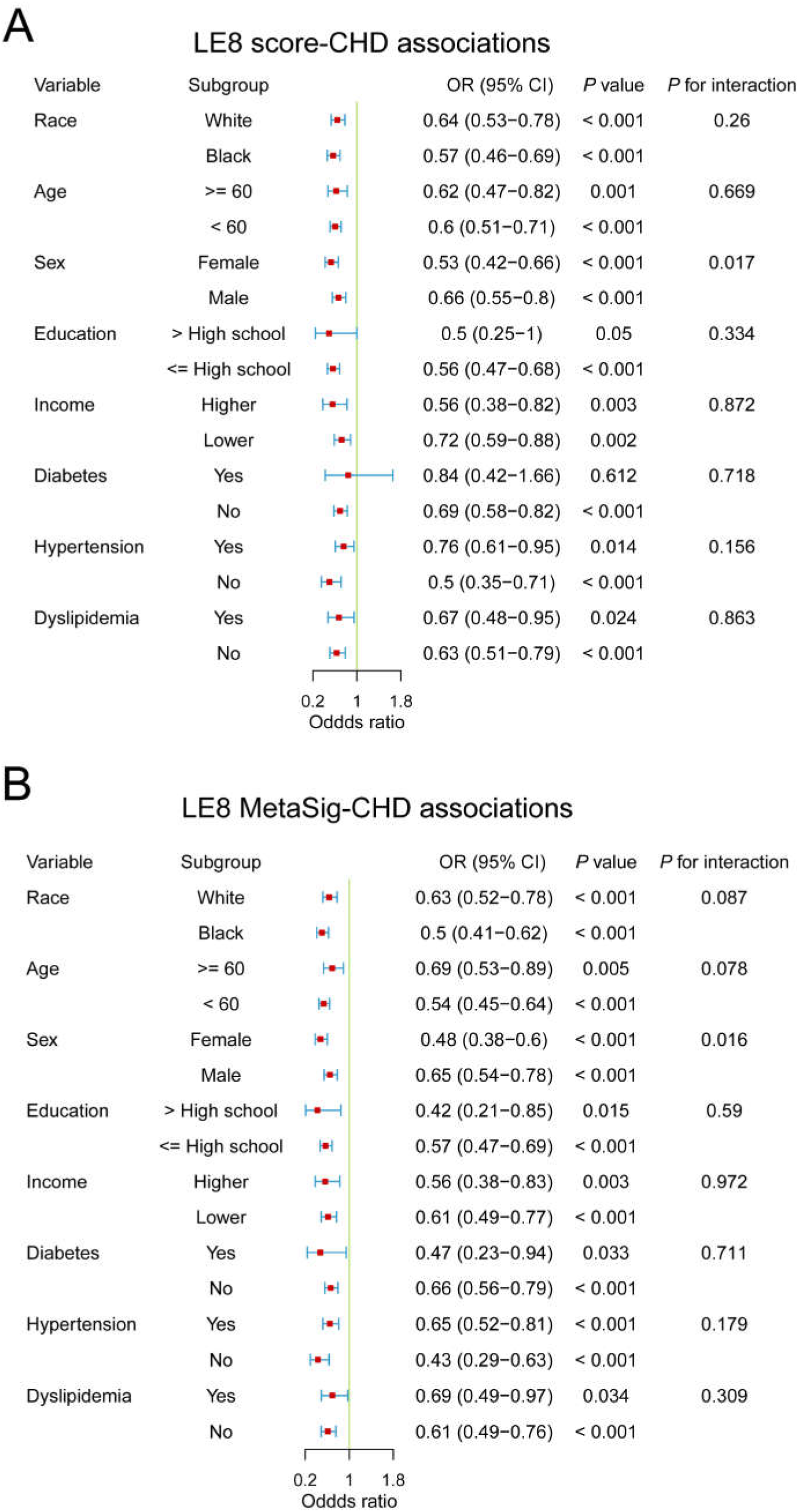
Subgroup analyses for the associations of LE8 score and its metabolite signature with risk of CHD in SCCS. **(A)** Subgroup analysis for the association between LE8 score and risk of CHD. **(B)** Subgroup analysis for the association between MetaSig and risk of CHD. Conditional logistic regression models were used, adjusted for potential confounders. For income, lower income denotes low income and higher income denotes middle and high income. **Abbreviations:** SCCS, Southern Community Cohort Study; MetaSig, metabolite signature; CHD, coronary heart disease; LE8, Life’s Essential 8; OR, odds ratio; CI, confidence interval.

The association of MetaSig with incident CHD was replicated in SWMHS (**Table 2)**, with OR (95% CI) = 0.57 (0.46-0.69) and 0.69 (0.55-0.86) after further adjusting for LE8 score (both *P*<0.001). The MetaSig also mediated a considerable portion of the LE8-CHD association in SWMHS [27.4% (10%-47%); *P*_mediation_<0.001; **Fig. 2E**].

### Metabolite signatures of health behaviors and health factors and associations with CHD

We further identified MetaSigs for health behaviors (**Fig. S2A**) and health factors in SCCS (**Fig. S3A**; full lists of metabolites are shown in **Table S3**), which showed strong correlations with health behaviors score (*r* = 0.59, *P*<0.001; **Fig. S2B**) and with health factors score (*r* = 0.76, *P*<0.001; **Fig. S3B**). Significant correlations were also found among participant subgroups (**Table S4**), suggesting the robustness of identified metabolite signatures for health behaviors and health factors. In addition, there were significant inverse associations of health behaviors score, health factors score, and their related signatures with risk of CHD (all *P*<0.001; **Table 2**). Specifically, standardized OR (95% CI) was 0.73 (0.63-0.85) for health behaviors MetaSig and 0.57 (0.49-0.66) for health factors MetaSig. Similarly, metabolites mediated large portions of the health behaviors-CHD association [43.9% (13.9%-101%), *P*=0.004; **Fig. S2D**] and health factors-CHD association [53.2% (24.8%-89%), *P*_mediation_<0.001; **Fig. S3D**]. Further, all results on health behaviors MetaSig and health factors MetaSig were replicated in SWMHS (**Table 2**, **Fig. S2**, and **Fig. S3**).

## Discussion

Leveraging untargeted plasma metabolites data in a nested case-control study among low-income Black and White Americans, we identified a metabolite signature that could reflect LE8 score and was associated with incident CHD, even after adjusting for LE8 and among participants with varied sociodemographic and metabolic health status, suggesting that circulating metabolite profiling may be used to help assess LE8 alignment across diverse populations and offer additional information on CVH (e.g., inter-individual metabolic phenotypes related to LE8). We also identified MetaSigs for health behaviors and health factors and found consistent results showing that circulating metabolites could reflect the alignment with those recommendations and underlying metabolic phenotypes, which were further linked to incident CHD across diverse populations. All the results were further replicated in another nested case-control study of CHD among Chinese adults. Our findings demonstrate the potential utility of blood metabolomics to improve the assessment of LE8 and its underlying metabolic variations that are linked to incident CHD among sociodemographically diverse populations, towards advancing precision medicine and addressing disparities in CVH.

LE8 is the American Heart Association’s updated and enhanced guideline to measure and promote CVH for individuals and populations^3^. The beneficial associations of following the LE8 with lower risks of CHD, CVD, and related mortality have been demonstrated in recent studies^4–7^. However, multi-racial/ethnic populations with low SES remain underrepresented in research studies, even though they have persistently experienced worse CVH and CVD outcomes, as well as systemic disadvantages to improve CVH, than White and middle-class Americans^2, 41–43^. Leveraging resources from SCCS, a large cohort of predominantly low-income Black and White Americans (in the present study: ∼65% with household income <$15,000/y and ∼95% with household income <$25,000/y), our study assessed CVH based on LE8 and evaluated the association of LE8 score with incident CHD. We found that higher LE8 score (per SD increase) was associated with ∼40-50% lower risk of CHD among Black Americans and individuals with low SES. While LE8 provides a comprehensive approach to quantify CVH, its assessment involves a series of procedures such as questionnaires, anthropometric and blood pressure measures, and blood draw. Particularly, health behaviors (diet, physical activity, smoking, and sleep) are usually assessed by questionnaires, which are time-consuming and prone to measurement errors and low compliance (particularly in the clinical setting). Also, LE8 score cannot capture varied individual metabolic responses to lifestyle exposures. Hence, we incorporated untargeted plasma metabolomics data, and for the first time, identified a robust metabolite signature of LE8 and then examined its association with incident CHD.

Metabolomics has been demonstrated as a powerful tool for improving exposure assessment and identifying potential novel biomarkers and mechanistic pathways in population studies, given its high-throughput characterization of thousands of metabolites in a small amount of biological samples^44^. Our study provides new evidence that plasma metabolite profiling may provide objective and comprehensive measures of LE8 and CVH among racially and geographically diverse populations. The identified metabolite signature may complement LE8 scores, improve the precision to stratify individuals with different future CHD risk, and potentially facilitate personalized CHD prevention strategies.

Our identified MetaSig consists of metabolites reflecting participants’ alignment with LE8 health behaviors and metabolic health status, majority of which are lipids and amino acids, and many of them have been linked to diet^12–16^, physical activity^17–19^, smoking^20–22^, sleep^23–26^, obesity^27, 28, 45^, or composite lifestyles scores^46–49^ in previous studies. For example, (2,4 or 2,5)-dimethylphenol sulfate, tartronate, and ethyl beta-glucopyranoside are derived from plant-based foods; cotinine is the major metabolite of nicotine from tobacco smoking; cholesterol, sphingomyelin, cortisol, and 1-palmitoleoylglycerolare are related to blood lipids; mannose, metformin, and fructosyllysine are related to prevalent diabetes and blood glucose. Particularly, drug metabolites including hydrochlorothiazide and metformin reflect antihypertensive and antidiabetic medications defined in LE8. Nevertheless, the associations of MetaSig with LE8 score and incident CHD were consistent among participants with or without history of hypertension or diabetes (**Fig. 3** and **Table S4**), and the MetaSig-CHD association did not change after excluding those drug metabolites from the MetaSig [OR (95% CI)=0.55 (0.48-0.64); *P*<0.001]. Moreover, excluding 29 unknown metabolites (X-) from the MetaSig also did not change the MetaSig-CHD association [OR (95% CI)=0.58 (0.50-0.66); *P*<0.001]. Notably, the MetaSig also contains microbial metabolites, e.g., maltotetraose, anthranilate, indolebutyrate, and bile acids (taurohyocholate, glycodeoxycholate 3-sulfate, 3b-hydroxy-5-cholenoic acid, and glycohyocholate), suggesting the role of gut microbiome in host’s CVH, which cannot be captured by questionnaires or measurements of glucose, cholesterol, or blood pressure. Moreover, several metabolic pathways related to CVH and CVD development were highlighted. For example, anthranilate, indolebutyrate, picolinate, and serotonin are members of tryptophan metabolism pathway^50–52^; taurohyocholate, glycodeoxycholate 3-sulfate, 3b-hydroxy-5-cholenoic acid, and glycohyocholate belong to secondary bile acid metabolism pathway^53, 54^; alpha-tocopherol, delta-tocopherol, and gamma-tocopherol/beta-tocopherol are vitamin E derivatives through tocopherol metabolism pathway^55, 56^.

Importantly, the MetaSig was related to future CHD risk regardless of participants’ age, sex, race, SES, metabolic disease history, and even after adjustment for LE8. Further analyses indicated that circulating metabolites could play a substantial mediating role linking LE8 and reduced CHD risk. Moreover, our findings were replicated in a racially and geographically different population, suggesting external validity and potential generalizability of our findings. Taken together, our findings demonstrated that circulating metabolites could complement LE8 to improve the precision of CVH assessment and predict CHD risk among sociodemographically and geographically diverse populations.

To our knowledge, this is the first study that assessed the LE8 score, constructed its metabolite signature, and associated LE8 score and its MetaSig with incident CHD in Black and White Americans with low SES. Besides its novelty and inclusion of populations facing socioeconomic challenges and health disparities, other strengths of our study include its prospective design, comprehensive profiling of >1500 blood metabolites for a broad coverage and improved ability to construct metabolite signature for LE8, and robustness of results across populations with different sociodemographic and health status. Meanwhile, several limitations of our current study need to be acknowledged. First, as MetaSig of LE8 was identified using cross-sectional data from baseline blood samples, we cannot be certain as to the directionality of LE8-MetaSig association, and mediation analysis assumed that LE8 score preceded MetaSig. Although several population- or animal-based studies have shown the causal effects of LE8 components on blood metabolites^21, 26, 57–60^, given that blood metabolites might precede some LE8 components, the longitudinal association between LE8 adherence and circulating metabolites should be investigated. Second, given the observational nature of our study, the causality is unable to be confirmed. However, the prospective design reduces the concern of reverse causation for the LE8/MetaSig-CHD association. Third, we cannot rule out the influence of residual confounding on the LE8/MetaSig-CHD association, although we have adjusted for and stratified by major CHD risk factors. Fourth, the concentrations of fasting glucose and HbA1c and SBP and DBP were not measured in SSCS; thus, glucose score and blood pressure score were defined based on history of diabetes or hypertension and use of medications, which may influence the accuracy of LE8 score. Finally, the nested case-control design may overestimate the predictive ability of the LE8 score and its MetaSig. Therefore, our results should be further validated in other prospective cohort studies.

In summary, our study assessed the LE8 score and identified MetaSig of LE8 among low-income Black and White Americans. We found that both LE8 score and its MetaSig were inversely associated with risk of CHD, consistently among participants with varied sociodemographic and metabolic health status. Our identified metabolite signature may provide an objective and comprehensive measure of LE8 and its metabolic underpinning, which may help improve the precision of CVH assessment and facilitate more effective and personalized CHD prevention strategies in diverse populations. Further examination of our identified metabolites may improve understanding of biological mechanisms as how following LE8 benefits CHD prevention.

## Data Availability

All data produced are available online at https://www.southerncommunitystudy.org/ for Southern Community Cohort Study and https://swhs-smhs.app.vumc.org/index.php for Shanghai Women's Health Study & Shanghai Men's Health Study.

## Acknowledgments

The authors thank the study participants of the SCCS and the SWMHS.

## References

1. Global burden of 369 diseases and injuries in 204 countries and territories, 1990-2019: a systematic analysis for the Global Burden of Disease Study 2019. Lancet 2020;396:1204–1222. doi: 10.1016/s0140-6736(20)30925-9

2. Tsao CW, Aday AW, Almarzooq ZI, et al. Heart Disease and Stroke Statistics-2023 Update: A Report From the American Heart Association. Circulation 2023. doi: 10.1161/cir.0000000000001123

3. Lloyd-Jones DM, Allen NB, Anderson CA, et al. Life’s essential 8: updating and enhancing the American Heart Association’s Construct of Cardiovascular Health: a presidential advisory from the American Heart Association. Circulation 2022;146:e18–e43. doi:

4. Wang X, Ma H, Li X, et al. Association of Cardiovascular Health With Life Expectancy Free of Cardiovascular Disease, Diabetes, Cancer, and Dementia in UK Adults. JAMA Intern Med 2023. doi: 10.1001/jamainternmed.2023.0015

5. Isiozor NM, Kunutsor SK, Voutilainen A, Laukkanen JA. Life’s Essential 8 and the risk of cardiovascular disease death and all-cause mortality in Finnish men. Eur J Prev Cardiol 2023. doi: 10.1093/eurjpc/zwad040

6. Jin C, Li J, Liu F, et al. Life’s Essential 8 and 10-Year and Lifetime Risk of Atherosclerotic Cardiovascular Disease in China. Am J Prev Med 2023. doi: 10.1016/j.amepre.2023.01.009

7. Yi J, Wang L, Guo X, Ren X. Association of Life’s Essential 8 with all-cause and cardiovascular mortality among US adults: A prospective cohort study from the NHANES 2005-2014. Nutr Metab Cardiovasc Dis 2023. doi: 10.1016/j.numecd.2023.01.021

8. Xanthakis V, Enserro DM, Murabito JM, et al. Ideal cardiovascular health: associations with biomarkers and subclinical disease and impact on incidence of cardiovascular disease in the Framingham Offspring Study. Circulation 2014;130:1676–1683. doi:

9. Gaye B, Tafflet M, Arveiler D, et al. Ideal cardiovascular health and incident cardiovascular disease: heterogeneity across event subtypes and mediating effect of blood biomarkers: the prime study. Journal of the American Heart Association 2017;6:e006389. doi:

10. Després JP. Predicting longevity using metabolomics: a novel tool for precision lifestyle medicine? Nat Rev Cardiol 2020;17:67–68. doi: 10.1038/s41569-019-0310-2

11. Clish CB. Metabolomics: an emerging but powerful tool for precision medicine. Molecular Case Studies 2015;1:a000588. doi:

12. Li J, Guasch-Ferré M, Chung W, et al. The Mediterranean diet, plasma metabolome, and cardiovascular disease risk. Eur Heart J 2020;41:2645–2656. doi: 10.1093/eurheartj/ehaa209

13. Chen L, Zhernakova DV, Kurilshikov A, et al. Influence of the microbiome, diet and genetics on inter-individual variation in the human plasma metabolome. Nat Med 2022. doi: 10.1038/s41591-022-02014-8

14. Wang F, Baden MY, Guasch-Ferré M, et al. Plasma metabolite profiles related to plant-based diets and the risk of type 2 diabetes. Diabetologia 2022;65:1119–1132. doi: 10.1007/s00125-022-05692-8

15. Chen GC, Chai JC, Xing J, et al. Healthful eating patterns, serum metabolite profile and risk of diabetes in a population-based prospective study of US Hispanics/Latinos. Diabetologia 2022;65:1133–1144. doi: 10.1007/s00125-022-05690-w

16. Shah RV, Steffen LM, Nayor M, et al. Dietary metabolic signatures and cardiometabolic risk. European Heart Journal 2022. doi:

17. Kemppainen SM, Fernandes Silva L, Lankinen MA, Schwab U, Laakso M. Metabolite Signature of Physical Activity and the Risk of Type 2 Diabetes in 7271 Men. Metabolites 2022;12. doi: 10.3390/metabo12010069

18. Kujala UM, Mäkinen VP, Heinonen I, et al. Long-term leisure-time physical activity and serum metabolome. Circulation 2013;127:340–348. doi: 10.1161/circulationaha.112.105551

19. Ding M, Zeleznik OA, Guasch-Ferre M, et al. Metabolome-Wide Association Study of the Relationship Between Habitual Physical Activity and Plasma Metabolite Levels. Am J Epidemiol 2019;188:1932–1943. doi: 10.1093/aje/kwz171

20. Tan Y, Barr DB, Ryan PB, et al. High-resolution metabolomics of exposure to tobacco smoke during pregnancy and adverse birth outcomes in the Atlanta African American maternal-child cohort. Environ Pollut 2022;292:118361. doi: 10.1016/j.envpol.2021.118361

21. Xu T, Holzapfel C, Dong X, et al. Effects of smoking and smoking cessation on human serum metabolite profile: results from the KORA cohort study. BMC Med 2013;11:60. doi: 10.1186/1741-7015-11-60

22. Gu F, Derkach A, Freedman ND, et al. Cigarette smoking behaviour and blood metabolomics. Int J Epidemiol 2016;45:1421–1432. doi: 10.1093/ije/dyv330

23. Bos MM, Goulding NJ, Lee MA, et al. Investigating the relationships between unfavourable habitual sleep and metabolomic traits: evidence from multi-cohort multivariable regression and Mendelian randomization analyses. BMC Med 2021;19:69. doi: 10.1186/s12916-021-01939-0

24. Fritz J, Huang T, Depner CM, et al. Sleep duration, plasma metabolites, and obesity and diabetes: A metabolome-wide association study in US women. Sleep 2022. doi: 10.1093/sleep/zsac226

25. Topriceanu CC, Tillin T, Chaturvedi N, Joshi R, Garfield V. The association between plasma metabolites and sleep quality in the Southall and Brent Revisited (SABRE) Study: A cross-sectional analysis. J Sleep Res 2021;30:e13245. doi: 10.1111/jsr.13245

26. Huang T, Zeleznik OA, Poole EM, et al. Habitual sleep quality, plasma metabolites and risk of coronary heart disease in post-menopausal women. Int J Epidemiol 2019;48:1262–1274. doi: 10.1093/ije/dyy234

27. Pan XF, Chen ZZ, Wang TJ, et al. Plasma metabolomic signatures of obesity and risk of type 2 diabetes. Obesity (Silver Spring) 2022. doi: 10.1002/oby.23549

28. Ottosson F, Smith E, Ericson U, et al. Metabolome-Defined Obesity and the Risk of Future Type 2 Diabetes and Mortality. Diabetes Care 2022;45:1260–1267. doi: 10.2337/dc21-2402

29. Signorello LB, Hargreaves MK, Blot WJ. The Southern Community Cohort Study: investigating health disparities. J Health Care Poor Underserved 2010;21:26–37. doi: 10.1353/hpu.0.0245

30. Zheng W, Chow WH, Yang G, et al. The Shanghai Women’s Health Study: rationale, study design, and baseline characteristics. Am J Epidemiol 2005;162:1123–1131. doi: 10.1093/aje/kwi322

31. Shu XO, Li H, Yang G, et al. Cohort Profile: The Shanghai Men’s Health Study. Int J Epidemiol 2015;44:810–818. doi: 10.1093/ije/dyv013

32. Lloyd-Jones DM, Ning H, Labarthe D, et al. Status of Cardiovascular Health in US Adults and Children Using the American Heart Association’s New “Life’s Essential 8” Metrics: Prevalence Estimates From the National Health and Nutrition Examination Survey (NHANES), 2013 Through 2018. Circulation 2022;146:822–835. doi: 10.1161/circulationaha.122.060911

33. Mellen PB, Gao SK, Vitolins MZ, Goff DC. Deteriorating dietary habits among adults with hypertension: DASH dietary accordance, NHANES 1988-1994 and 1999-2004. Archives of internal medicine 2008;168:308–314. doi:

34. Chang RS, Xu M, Brown SH, et al. Relation of the Dietary Approaches to Stop Hypertension Dietary Pattern to Heart Failure Risk and Socioeconomic Status (from the Southern Community Cohort Study). Am J Cardiol 2022;169:71–77. doi: 10.1016/j.amjcard.2021.12.043

35. Yu D, Zhang X, Xiang YB, et al. Adherence to dietary guidelines and mortality: a report from prospective cohort studies of 134,000 Chinese adults in urban Shanghai. Am J Clin Nutr 2014;100:693–700. doi: 10.3945/ajcn.113.079194

36. Evans AM, DeHaven CD, Barrett T, Mitchell M, Milgram E. Integrated, nontargeted ultrahigh performance liquid chromatography/electrospray ionization tandem mass spectrometry platform for the identification and relative quantification of the small-molecule complement of biological systems. Anal Chem 2009;81:6656–6667. doi: 10.1021/ac901536h

37. Zou H, Hastie T. Regularization and variable selection via the elastic net. Journal of the royal statistical society: series B (statistical methodology) 2005;67:301–320. doi:

38. Kuhn M. Building predictive models in R using the caret package. Journal of statistical software 2008;28:1–26. doi:

39. Friedman J, Hastie T, Tibshirani R. Regularization paths for generalized linear models via coordinate descent. Journal of statistical software 2010;33:1. doi:

40. Tingley D, Yamamoto T, Hirose K, Keele L, Imai K. Mediation: R package for causal mediation analysis. 2014. doi:

41. Huffman MD, Capewell S, Ning H, et al. Cardiovascular health behavior and health factor changes (1988-2008) and projections to 2020: results from the National Health and Nutrition Examination Surveys. Circulation 2012;125:2595–2602. doi: 10.1161/circulationaha.111.070722

42. Pilkerton CS, Singh SS, Bias TK, Frisbee SJ. Changes in Cardiovascular Health in the United States, 2003-2011. J Am Heart Assoc 2015;4:e001650. doi: 10.1161/jaha.114.001650

43. Haynes N, Kaur A, Swain J, Joseph JJ, Brewer LC. Community-Based Participatory Research to Improve Cardiovascular Health Among US Racial and Ethnic Minority Groups. Curr Epidemiol Rep 2022;9:212–221. doi: 10.1007/s40471-022-00298-5

44. Wishart DS. Emerging applications of metabolomics in drug discovery and precision medicine. Nature reviews Drug discovery 2016;15:473–484. doi:

45. Cirulli ET, Guo L, Leon Swisher C, et al. Profound Perturbation of the Metabolome in Obesity Is Associated with Health Risk. Cell Metab 2019;29:488–500.e482. doi: 10.1016/j.cmet.2018.09.022

46. Delgado-Velandia M, Gonzalez-Marrachelli V, Domingo-Relloso A, et al. Healthy lifestyle, metabolomics and incident type 2 diabetes in a population-based cohort from Spain. Int J Behav Nutr Phys Act 2022;19:8. doi: 10.1186/s12966-021-01219-3

47. Rothwell JA, Murphy N, Bešević J, et al. Metabolic Signatures of Healthy Lifestyle Patterns and Colorectal Cancer Risk in a European Cohort. Clin Gastroenterol Hepatol 2022;20:e1061–e1082. doi: 10.1016/j.cgh.2020.11.045

48. Assi N, Gunter MJ, Thomas DC, et al. Metabolic signature of healthy lifestyle and its relation with risk of hepatocellular carcinoma in a large European cohort. Am J Clin Nutr 2018;108:117–126. doi: 10.1093/ajcn/nqy074

49. Si J, Li J, Yu C, et al. Improved lipidomic profile mediates the effects of adherence to healthy lifestyles on coronary heart disease. Elife 2021;10. doi: 10.7554/eLife.60999

50. Wirleitner B, Rudzite V, Neurauter G, et al. Immune activation and degradation of tryptophan in coronary heart disease. European journal of clinical investigation 2003;33:550–554. doi:

51. Mangge H, Stelzer I, Reininghaus E, et al. Disturbed tryptophan metabolism in cardiovascular disease. Current medicinal chemistry 2014;21:1931–1937. doi:

52. Liu G, Chen S, Zhong J, Teng K, Yin Y. Crosstalk between tryptophan metabolism and cardiovascular disease, mechanisms, and therapeutic implications. Oxidative Medicine and Cellular Longevity 2017;2017. doi:

53. Fan Y, Pedersen O. Gut microbiota in human metabolic health and disease. Nat Rev Microbiol 2021;19:55–71. doi: 10.1038/s41579-020-0433-9

54. Agus A, Clément K, Sokol H. Gut microbiota-derived metabolites as central regulators in metabolic disorders. Gut 2021;70:1174–1182. doi: 10.1136/gutjnl-2020-323071

55. Dietrich M, Traber MG, Jacques PF, et al. Does γ-tocopherol play a role in the primary prevention of heart disease and cancer? A review. Journal of the American College of Nutrition 2006;25:292–299. doi:

56. Mathur P, Ding Z, Saldeen T, Mehta JL. Tocopherols in the prevention and treatment of atherosclerosis and related cardiovascular disease. Clinical cardiology 2015;38:570–576. doi:

57. Würtz P, Wang Q, Kangas AJ, et al. Metabolic signatures of adiposity in young adults: Mendelian randomization analysis and effects of weight change. PLoS Med 2014;11:e1001765. doi: 10.1371/journal.pmed.1001765

58. Lauterbach MA, Latz E, Christ A. Metabolomic Profiling Reveals Distinct and Mutual Effects of Diet and Inflammation in Shaping Systemic Metabolism in Ldlr(-/-) Mice. Metabolites 2020;10. doi: 10.3390/metabo10090336

59. Sato S, Dyar KA, Treebak JT, et al. Atlas of exercise metabolism reveals time-dependent signatures of metabolic homeostasis. Cell Metabolism 2022;34:329–345. e328. doi:

60. Cruickshank-Quinn CI, Mahaffey S, Justice MJ, et al. Transient and persistent metabolomic changes in plasma following chronic cigarette smoke exposure in a mouse model. PloS one 2014;9:e101855. doi:

