## Supplementary materials for "Metabolite Signature of Life’s Essential 8 and Risk of Coronary Heart Disease among Low-Income Black and White Americans"

### **Table S1. Measurement of alignment with Life’s Essential 8 components**

| LE8 components | Measurement | Scoring | | | | | | | |
| --- | --- | --- | --- | --- | --- | --- | --- | --- | --- |
|  |  | SCCS | | | | | SWMHS | | |
| Diet score | DASH score | Points | | | | Quantile | Points | Quantile | |
|  |  | 100 | | | | ≥95th percentile | 100 | ≥95th percentile | |
|  |  | 80 | | | | 75th-94th percentile | 80 | 75th-94th percentile | |
|  |  | 50 | | | | 50th-74th percentile | 50 | 50th-74th percentile | |
|  |  | 25 | | | | 25th-49th percentile | 25 | 25th-49th percentile | |
|  |  | 0 | | | | 1th-24th percentile | 0 | 1th-24th percentile | |
| Physical activity score | Self-reported total minutes of leisure-time moderate and vigorous physical activity per week | Points | | | | Minutes | Points | Minutes | |
|  |  | 100 | | | | ≥150 | 100 | ≥150 | |
|  |  | 90 | | | | 120-149 | 90 | 120-149 | |
|  |  | 80 | | | | 90-119 | 80 | 90-119 | |
|  |  | 60 | | | | 60-89 | 60 | 60-89 | |
|  |  | 40 | | | | 30-59 | 40 | 30-59 | |
|  |  | 20 | | | | 1-29 | 20 | 1-29 | |
|  |  | 0 | | | | 0 | 0 | 0 | |
| Smoking score | Tobacco smoking and secondhand smoke exposure | Points | | | | Status | Points | Status | |
|  |  | 100 | | | | Never smoker | 100 | Never smoker | |
|  |  | 75 | | | | Former smoker, quit ≥5y | 75 | Former smoker, quit ≥5y | |
|  |  | 50 | | | | Former smoker, quit 1-<5y | 50 | Former smoker, quit 1-<5y | |
|  |  | 25 | | | | Former smoker, quit <1y | 25 | Former smoker, quit <1y | |
|  |  | 0 | | | | Current smoker | 0 | Current smoker | |
|  |  | Subtract 20 points (unless score is 0) for exposing to secondhand smoking | | | | | Subtract 20 points (unless score is 0) for exposing to secondhand smoking | | |
| Sleep score | Average self-reported sleep hours per day | Points | | | | Level | Points | Levels | |
|  |  | 100 | | | | 7-<9 | 100 | 7-<9 | |
|  |  | 90 | | | | 9-<10 | 90 | 9-<10 | |
|  |  | 70 | | | | 6-<7 | 70 | 6-<7 | |
|  |  | 40 | | | | 5-<6 or ≥10 | 40 | 5-<6 or ≥10 | |
|  |  | 20 | | | | 4-<5 | 20 | 4-<5 | |
|  |  | 0 | | | | <4 | 0 | <4 | |
| BMI score | Weight (kg)/height (m)^2^ | Points | | | Level | | Points | Level | |
|  |  | 100 | | | <25 | | 100 | <23 | |
|  |  | 70 | | | 25.0-29.9 | | 75 | 23.0-24.9 | |
|  |  | 30 | | | 30.0-34.9 | | 50 | 25.0-29.9 | |
|  |  | 15 | | | 35.0-39.9 | | 25 | 30.0-34.9 | |
|  |  | 0 | | | ≥40.0 | | 0 | ≥35.0 | |
| Lipids score | Non-HDL cholesterol (mg/dL) | Points | | Level | | | Points | Level | |
|  |  | 100 | | <130 | | | 100 | <130 | |
|  |  | 60 | | 130-159 | | | 60 | 130-159 | |
|  |  | 40 | | 160-189 | | | 40 | 160-189 | |
|  |  | 20 | | 190-219 | | | 20 | 190-219 | |
|  |  | 0 | | ≥220 | | | 0 | ≥220 | |
|  |  | If drug-treated level, subtract 20 points (unless score is 0) | | | | | If drug-treated level, subtract 20 points (unless score is 0) | | |
| Glucose score | T2D status and glucose abundance measured by untargeted metabolomic profiling | Points | Level | | | | Points | Level | |
|  |  | Without diabetes | | | | | Without diabetes | | |
|  |  | 100 | Glucose <50th percentile | | | | 100 | Glucose <50th percentile | |
|  |  | 60 | Glucose ≥50th percentile | | | | 60 | Glucose ≥50th percentile | |
|  |  | With diabetes | | | | | With diabetes | | |
|  |  | 40 | Glucose abundance <20th percentile | | | | 40 | Glucose abundance <20th percentile | |
|  |  | 30 | Glucose abundance 20th-<40th percentile | | | | 30 | Glucose abundance 20th-<40th percentile | |
|  |  | 20 | Glucose abundance 40th-<60th percentile | | | | 20 | Glucose abundance 40th-<60th percentile | |
|  |  | 10 | Glucose abundance 60th-<80th percentile | | | | 10 | Glucose abundance 60th-<80th percentile | |
|  |  | 0 | Glucose abundance ≥80th percentile | | | | 0 | Glucose abundance ≥80th percentile | |
| Blood pressure score | SCCS: hypertension status and number of medications; SWMHS: SBP (mmHg) and DBP (mmHg) | Points | Level | | | | Points | | Level |
|  |  | 100 | Without hypertension | | | | 100 | | SBP<120 and DBP<80 |
|  |  | With hypertension | | | | | 75 | | SBP 120-<130 and DBP<80 |
|  |  | 50 | Without taking medications | | | | 50 | | SBP 130-<140 or DBP 80-<90 |
|  |  | 25 | Taking only 1 medication | | | | 25 | | SBP 140-<160 or DBP 90-<100 |
|  |  | 0 | Taking >1 medication | | | | 0 | | SBP≥160 or DBP≥100 |
|  |  |  |  | | | | Subtract 20 points if participant receive anti-hypertension treatment (minimal score is 0) | | |

### **Table S2. Characteristics of study participants in Shanghai Women’s and Men’s Health Studies**

|  | CHD (*N*=299) | Control (*N*=299) |
| --- | --- | --- |
| Age, years | 61.5 (8.3) | 61.4 (8.3) |
| Male, *n* (%) | 149 (49.8) | 149 (49.8) |
| Education, *n* (%) |  |  |
| Less than high school | 181 (60.5) | 184 (61.5) |
| Completed high school | 67 (22.4) | 61 (20.4) |
| Vocational school or some college | 29 (9.7) | 22 (7.4) |
| College or graduate school | 22 (7.4) | 32 (10.7) |
| Income, *n* (%)* |  |  |
| Low | 59 (19.7) | 54 (18.1) |
| Middle | 224 (74.9) | 231 (77.3) |
| High | 16 (5.4) | 14 (4.7) |
| Alcohol intake, *n* (%)** |  |  |
| None | 258 (86.3) | 255 (85.3) |
| Moderate | 22 (7.4) | 31 (10.4) |
| Heavy | 19 (6.4) | 13 (4.3) |
| Family history of CHD, *n* (%) | 43 (14.4) | 33 (11.0) |
| History of diabetes, *n* (%) | 58 (19.4) | 25 (8.4) |
| History of dyslipidemia, *n* (%) | 34 (11.4) | 22 (7.4) |
| History of hypertension, *n* (%) | 151 (50.5) | 101 (33.8) |
| LE8 score | 50.7 (12.0) | 57.2 (12.8) |
| LE8 score category, *n* (%)*** |  |  |
| High (80-100) | 1 (0.3) | 7 (2.3) |
| Moderate (50-79) | 160 (53.5) | 207 (69.2) |
| Low (0-49) | 138 (46.2) | 85 (28.4) |
| Health behaviors score | 53.6 (21.8) | 58.0 (22.3) |
| Health factors score | 47.8 (15.6) | 56.2 (15.7) |
| Diet score | 35.5 (30.4) | 44.0 (31.8) |
| Physical activity score | 44.6 (49.0) | 46.4 (48.9) |
| Smoking score | 61.4 (40.4) | 67.7 (38.2) |
| Sleep score | 79.9 (24.6) | 79.9 (25.0) |
| BMI score | 68.4 (25.8) | 73.2 (22.9) |
| Blood lipids score | 38.0 (31.9) | 48.3 (32.5) |
| Blood glucose score | 66.9 (30.2) | 76.4 (25.5) |
| Blood pressure score | 15.0 (27.2) | 25.6 (35.8) |

Data were mean (standard deviation) or *n* (%) as indicated.

*Annual income per capita <￥6,000, ￥6,000 to <￥10,000, and ≥￥10,000 for low, middle, and high levels of income, respectively in Chinese men and <￥4000, ￥4,000 to <￥8,000, and ≥￥8,000 for low, middle, and high levels of income, respectively in Chinese women.

**Alcohol intake was grouped as none, moderate (>0 to ≤2 drinks per day in men or >0 to ≤1 drink per day in women; 1 drink = 14 g ethanol), and heavy drinking (>2 drinks per day in men or >1 drink per day in women).

***The cutoffs were provided by the American Heart Association (Lloyd-Jones *et al.*, 2022).

Abbreviations: CHD, coronary heart disease; LE8, Life’s Essential 8; BMI, body mass index.

### **Fig. S1. Correlations between LE8 score and its individual component scores in SCCS (A) and SWMHS (B).** Values in the figures are *Spearman* correlation coefficients. Colors represent the extent of correlations.

Abbreviations: LE8, life’s Essential 8; BMI, body mass index. SCCS, Southern Community Cohort Study; SWMHS, Shanghai Women’s and Men’s Health Studies.

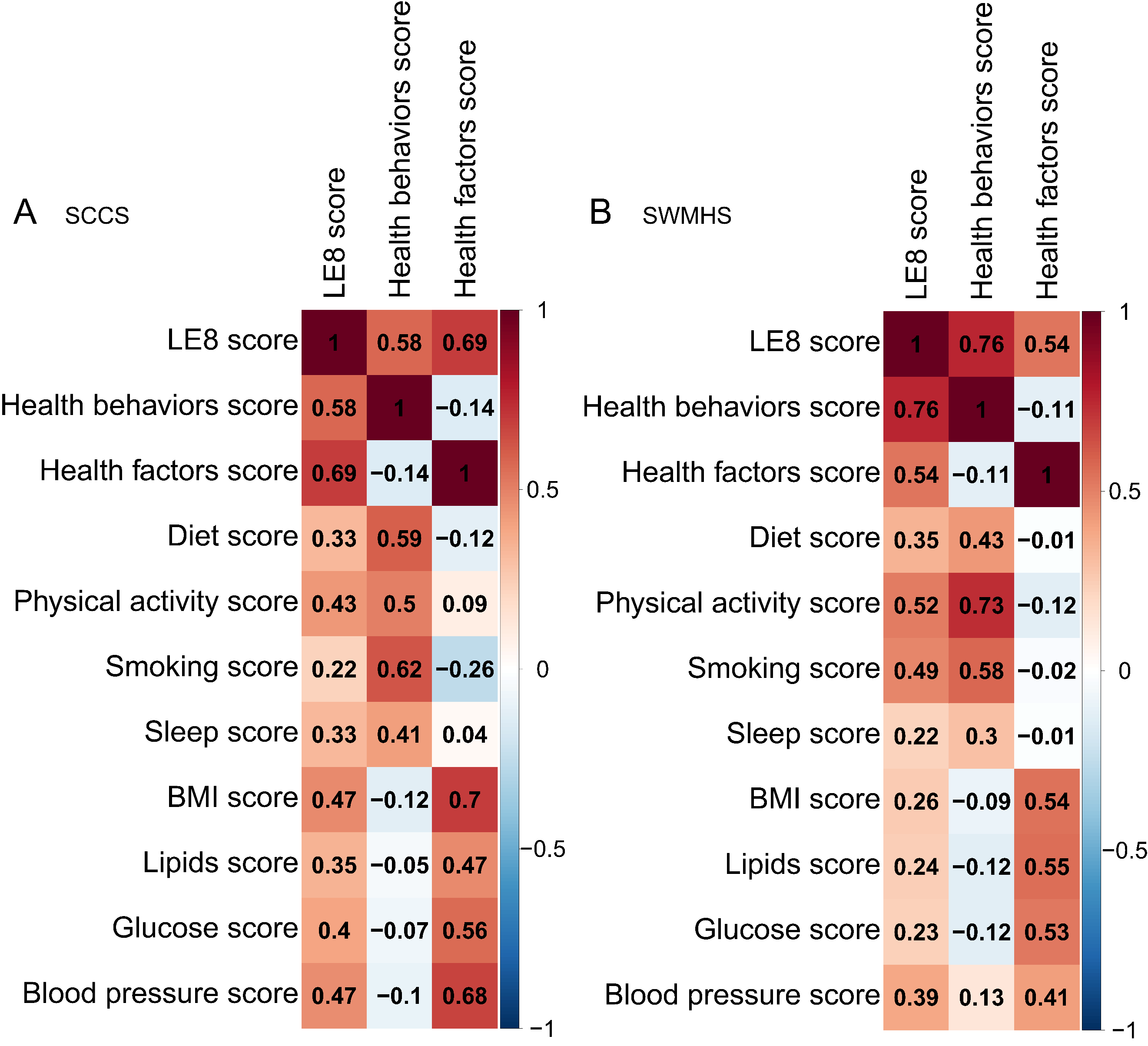

### **Fig. S2. The** **metabolite signature of health behaviors and its association with risk of CHD. (A)** Top 30 metabolites selected by elastic net regression in SCCS. Metabolites were ranked by the absolute value of regression coefficients. **(B)** *Spearman* correlation between MetaSig and health behaviors score in SCCS. **(C)** *Spearman* correlation between MetaSig and health behaviors score in SWMHS. **(D)** The mediation effect of MetaSig on the association between health behaviors score and risk of CHD in SCCS. **(E)** The mediation effect of MetaSig on the association between health behaviors score and risk of CHD in SWMHS.

**Abbreviations:** SCCS, Southern Community Cohort Study; SWMHS, Shanghai Women’s and Men’s Health Studies; MetaSig, metabolite signature; ACME, average causal mediation effects; ADE, average direct effects; CHD, coronary heart disease.

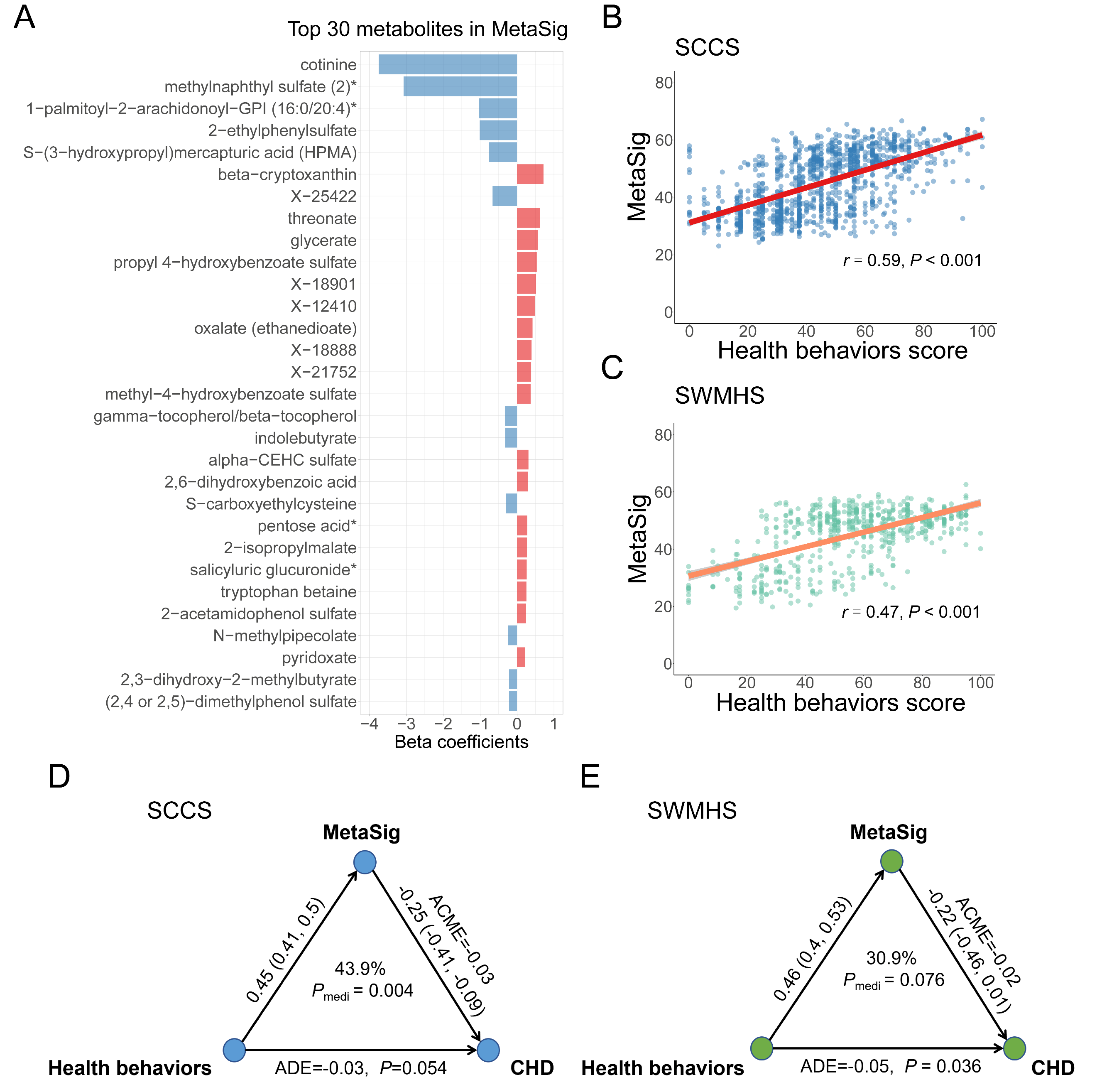

### **Fig. S3. The metabolite signature of health factors and its association with risk of CHD. (A)** Top 30 metabolites selected by elastic net regression in SCCS. Metabolites were ranked by the absolute value of regression coefficients. **(B)** *Spearman* correlation between MetaSig and health factors score in SCCS. **(C)** *Spearman* correlation between MetaSig and health factors score in SWMHS. **(D)** The mediation effect of MetaSig on the association between health factors score and risk of CHD in SCCS. **(E)** The mediation effect of MetaSig on the association between health factors score and risk of CHD in SWMHS.

**Abbreviations:** SCCS, Southern Community Cohort Study; SWMHS, Shanghai Women’s and Men’s Health Studies; MetaSig, metabolite signature; ACME, average causal mediation effects; ADE, average direct effects; CHD, coronary heart disease.

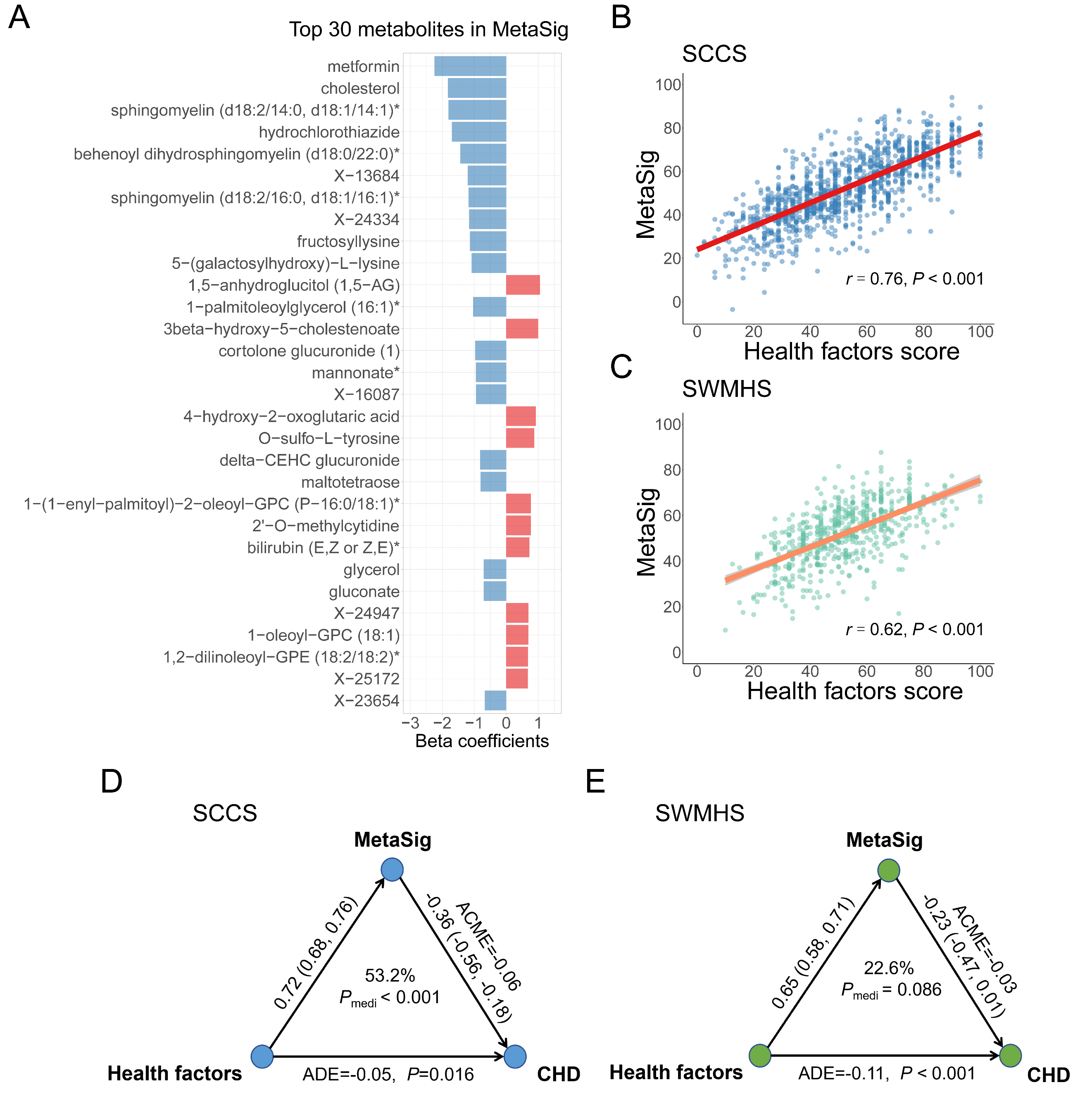
